# Present knowledge, attitude, practice, and fear level of Bangladeshi people towards covid-19 after a year of the pandemic situation: a web-based cross-sectional study

**DOI:** 10.1101/2021.07.19.21260721

**Authors:** Tahsin Ahmed Rupok, Sunandan Dey, Rashni Agarwala, Md. Nurnobi Islam, Bayezid Bostami

## Abstract

In the earlier phase of the pandemic situation, the governments of Bangladesh badly suffered to adhere their people to preventive measures probably due to less knowledge and attitude towards covid-19. To tackle the second wave of coronavirus, the governments again enforced an array of preventive measures, but still encountering the same problem after a year of the pandemic situation. In an attempt to find out the reasons behind this, our study aimed to assess the present knowledge, attitude, practice, and fear level of the people. A cross-sectional study was conducted from 15^th^ to 25^th^ April 2021. A total of 402 participants met all the inclusion criteria and were considered for performing all statistical analyses (Descriptive statistics, Mann-Whitney U test, Kruskal-Wallis H test, Multiple logistic regression, Spearman rank-order correlation). Out of 402 participants, more than 90% participants were students and all were adults aged 16 to 30. 84.6%, 65.7%, 54%, and 21.6% participants had more adequate knowledge, more positive attitude, more frequent practice, and moderate to high fear towards covid-19, respectively. Knowledge, attitude, practice, and fear were interrelated directly or indirectly. It was found knowledgeable participants were more likely to have more positive attitude (OR = 2.12, 95% CI = 1.14-3.95, P < 0.05) and very less fear (OR = 1.98, 95% CI = 1.02-3.82, P < 0.05). More positive attitude was found as a good predictor of more frequent practice (OR = 4.33, 95% CI = 2.66-7.04, P < 0.001), and very less fear had same negative impact on both attitude (OR = 0.48, 95% CI = 0.25-0.91, P < 0.05) and practice (OR = 0.48, 95% CI = 0.27-0.85, P < 0.05). Our findings reflect that knowledge level has elevated but attitude level subsided, and practice level stayed same as was in the earlier phase of pandemic and people are no longer panicked.

## Introduction

In Bangladesh, both infection and death rates have started to burgeon again from the second week of March 2021 with a small dip in late April, which has been further instigated due to the entrance of highly transmissible Delta variant of coronavirus in May this year (1). As of June 29, 2021, 7,666 new cases were detected with a positivity rate of 23.97, and 112 people lost their life (2). Infectious disease experts, Dr. Be-Nazir Ahmed, a former director at the Directorate General of Health Services (DGHS) predicted that if this Delta variant starts to escalate vehemently as it did in India and Nepal, people stay lackadaisical in maintaining public health hygiene, and governments continue to develop an ill-conceived plan of action and fail to ensure proper implementation of preventive measures, then, the number of cases detected per day could rise to 20,000 or even more to 40,000 in July. He also added if the country records over 10,000 cases for some weeks it will be difficult to control the situation as it does not have sufficient hospital beds, oxygen production, ICUs, doctors, nurses, and other facilities and equipment to take care of such a huge number of patients (3). Besides, there are still no acceptable treatments that can warrant a short period of hospitalized conditions. So, the government should take the necessary steps to prevent any grim situation. It is worth mentioning that prevention is the best and comparatively less costly method to the most effective protection against the virus. Only adequate preventive measures can keep the infection rate under control, which will truncate both the number of patients and death tallies. As a preventive measure, the government decided to go for countrywide strict lockdown again from 1^st^ July of this year but it is worth remembering that lockdowns are effective only if they are accompanied by other preventive measures (4,5). However, Bangladesh’s governments badly suffered to adhere their population to preventive measures in the earlier phase of the pandemic situation probably due to people’s less knowledge and less attitude to embrace the measures taken by the governments. A review of some previous studies conducted in Bangladesh reflected the truth that last year Bangladeshi people were comparatively less knowledgeable, had a less positive attitude toward covid-19, and as a consequence had less frequent practice, which was consistent with a study of Pakistan but inconsistent with China and Saudi Arabia. Indeed, there are also exceptions such as one article (Bangladesh 4) showed people (78.9%) had a comparatively more positive attitude towards covid-19. Moreover, another article (Bangladesh 5) reported that people had more frequent practice (94.3%) along with positive attitude (93.0%) (Figure 01) [Bangladesh 1 (6), Bangladesh 2 (7), Bangladesh 3 (8), Bangladesh 4 (9), Bangladesh 5 (10), Pakistan (11), Saudi Arabia (12), China (13)].

**Fig 1.**
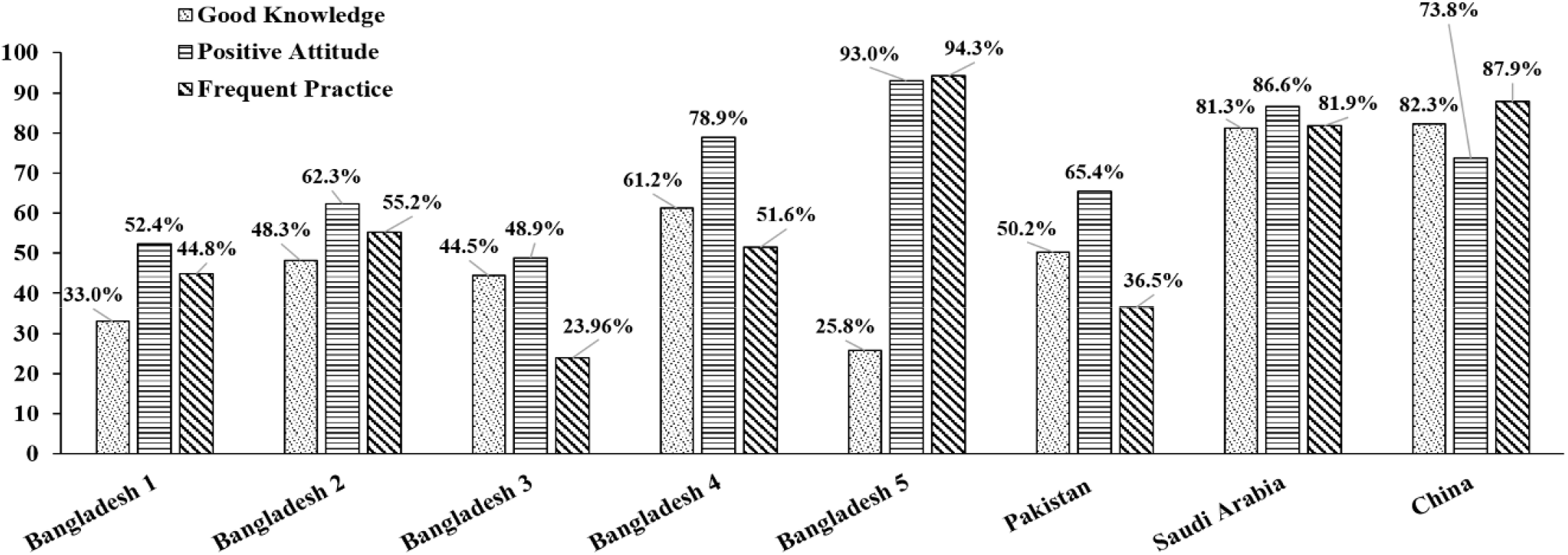
Diagram reflecting different countries and their peoples’ knowledge, attitude, and practice level towards covid-19.

**Fig 1.**
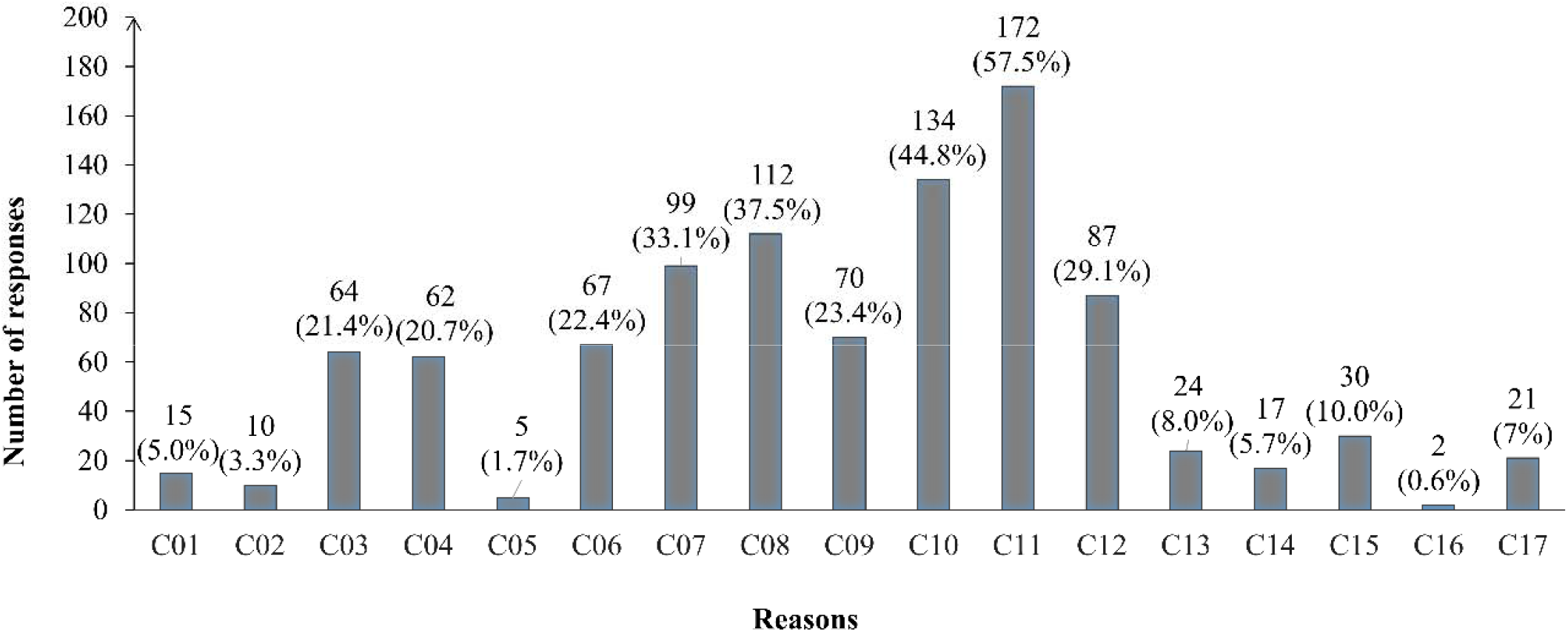
Graphical representation of reasons behind failure to maintain safety guidelines of covid-19. (Table 07) tabulated all the supportive information.

However, one thing to remember, adhering people to preventive measures is not straightforward, takes intensive efforts over a length of time as well as depends on what extent people can perceive the import of those measures. So, proper implementation of these preventive measures warrants adequate well-planned initiatives to scale up people’s knowledge and awareness towards covid-19 and its preventive measures, which will ultimately alter their behavior as well as attitude positively towards covid-19 preventive practice. Several studies published last year (2020) outlined that adequate knowledge regarding covid-19 is a potential predictor of attitude towards its preventive practice (8,14). To speak the truth, it is indefinite when the coronavirus will go away from the world. Therefore, people must be resilient to this new situation, learn all fundamental information regarding covid-19, and change their attitude positively towards practicing all safety guidelines stipulated by their government.

Our study aimed to bring up the present KAP level of Bangladeshi people after a year of the pandemic situation and how prepared they are to confront any future onslaught of covid-19. So, the objectives of this present study were-

‐ To explore the present knowledge, attitude, practice, and fear level of the Bangladeshi population and their associated potential factors.
‐ To find out whether knowledge, attitude, practice, and fear are related to each other.
‐ To find out the most probable reasons why people are digressed from maintaining preventive measures enforced by the governments.

## Materials and Methods

### Participants and Data recruitment procedure

This web-based cross-sectional survey was conducted on Bangladeshi people using online facilities amid the second wave of coronavirus in Bangladesh, continued for 10 days from 15^th^ to 25^th^ April in 2021. A structured questionnaire was prepared in a Google Form and a link was generated, shared with all authors and other volunteers who were instructed properly as to how well they can use this form to recruit data with adequate consent from the participants. A face-to-face community-based survey was not possible as the Bangladesh government implemented their second strict lockdown from 14^th^ April being one day ahead of our data collection initiated (15), all data were therefore assembled through online approaches. This survey included subjects aged 16 years old or above, being Bangladeshi residents, having internet access and ability to communicate, having no intellectual disability. Subjects living in foreign countries, having no or less interest to participate in were excluded from the survey. A total of 407 subjects successfully submitted their data and all completed the questionnaire adequately as they were recommended.

### Survey Instrument

The questionnaire prepared after a thorough review of some literature published last year had six sections. The first section with a brief delineation of the objectives of the survey and some instructions on how to complete the questionnaire properly entailed a question asked to take consent of the respondents. The second section embodied 7 questions asked to gather socio-demographic information of a participant. Socio-demographic information included gender, age, education, occupation, location, family type, Social status (family income) which was categorized as follows “Lower class” (<20,000 BDT), “Middle class” (20,000-40,000 BDT), “High class” (>40,000 BDT). The third section (Knowledge scale) had 12 questions where each question had three options “Right”, “Wrong”, and “Not sure”. The fourth section (Attribute scale) had 8 questions where each question had three options “Yes”, “No”, and “Maybe” except one question having “Not possible” instead “Maybe” in three options. The fifth section (Practice section) also had 8 questions where each question had three options “Yes”, “No”, and “Sometimes”. The sixth section (Fear scale) included a fear measuring tool which was reported as a valid and reliable instrument to assess covid-19 related fear among respondents (16). Each question had five Likert options. This section also had two extra questions. The first question having binary options “Yes”, “No” asked if or not they are maintaining preventative measures properly stipulated by the governments. The second question was asked to those who answered “No” to account for their reasons why they are aberrant than those who answered “Yes”. There were 17 possible responses and multiple responses could be selected.

As to scoring, in the knowledge scale, the correct answer (Right) was coded as 1 and the incorrect answer (Wrong/Don’t know) was coded as 0. The total score ranged from 0 to 12 where those who obtained 83% score or greater (≥ 10) were considered to have more accurate knowledge otherwise considered having less accurate knowledge.

In the attitude scale, four kinds of options were coded as follows, 0 (No/ Not possible for work), 1 (Maybe), and 2 (Yes). The total score ranged from 0 to 16.

Those who had an 81% score or greater (≥ 13) were considered to have a positive attitude otherwise considered to have a less positive attitude.

Three options of the practice scale were coded as follows, 0 (No), 1 (Sometimes), and 2 (Yes). The total score ranged from 0 to 16. Those who had an 81% score or greater (≥ 13) were considered to have more frequent practice otherwise considered having less frequent practice.

The fear scale had 7 items where each item had five Likert options which were coded as follows, 0 (Strongly disagree), 1 (Disagree), 2 (Neutral), 3 (Agree), 4 (Strongly Agree). The total score ranged from 0 to 28. Fear scores greater or equal to 21 were cut-points for high fear. Scores between 20-14 were cut points for moderate fear and scores equal to or less than 13 were cut points for less fear.

Reliability tests were performed using field data to determine the validity and reliability of those scales mentioned above. Cronbach’s alpha for knowledge, attitude, practice, and fear scale were 0.47, 0.61, 0.81, and 0.83. Due to a shortage of time, we could not conduct a pilot study.

### Ethical considerations

This study was conducted in accordance with the institutional Research Ethics and the declaration of Helsinki. Besides, this study had been approved by the ethical review committee of Rajshahi University. Anonymity and confidentiality were strictly maintained. All respondents gave full informed consent to participate and consent for their data to be used in the publication.

### Data Management and Analysis

All submitted data were automatically stored in a dynamic Microsoft excel sheet which was made available offline after completion of data collection for primary data processing including data duplication-checking, data cleaning, data coding, etc. In this stage, three data fallen out of inclusion criteria of age (below 15 years of old), and two data got duplicated were discard from the preliminary dataset. After accomplishment of data cleaning and coding, the final dataset having data of 402 respondents was entered into Statistical Package for the Social Science (SPSS) version 26.0 for further data processing and doing all statistical analysis which included descriptive statistics, Mann-Whitney U test, Kruskal Wallis H test, Multiple logistic regression, Spearman rank-order correlation. Non-parametric Mann-Whitney U test and Kruskal Wallis H test, and Spearman rank-order correlation were performed as data under continuous dependent variables were not normally distributed. To check that, besides visual inspection of histograms, Shapiro-Wilk and Kolmogorov-Smirnov tests were conducted. Both tests were statistically significant at 0.01 for those continuous dependent variables and histograms of them looked left-skewed, which confirmed well that data under these variables were not normally distributed. For all statistical analyses noted above, the statistical significance level was set to 0.05 (Alpha value).

## Results

### Demographic characteristics of the sample

This study considered a total of 402 participants for final statistical analysis. Among them majority were male (74.2%), studying in honors and masters (71.6%). More than 90% of participants were students while 37.8% were students and self-employed. About 75% and 56% of participants were from nuclear and lower class family respectively. About 36% of participants informed their location (for at least 3 months) village. This study inclined towards the adult population as all participants had ages of 16 to 30 years of old. (Table 01) tabulated detailed information about the socio-demographic characteristics of the respondents.

**Table 01.** Socio-demographic characteristics of the respondents.

| Characteristics |  | Frequency | Percentage (%) |
| --- | --- | --- | --- |
| Gender | Male | 300 | 74.2 |
|  | Female | 102 | 25.4 |
| Education | HSC | 114 | 28.4 |
|  | Honors/Masters/Above | 288 | 71.6 |
| Occupation | Student and self-employed | 152 | 37.8 |
|  | Student only | 230 | 57.2 |
|  | Jobs/Business | 20 | 5.0 |
| Family type | Nuclear | 304 | 75.6 |
|  | Extended | 98 | 24.4 |
| Social status | Lower class | 228 | 56.7 |
|  | Middle class | 135 | 33.6 |
|  | High class | 39 | 9.7 |
| Location (At least 3 months) | Village | 145 | 36.1 |
|  | Sub-district town | 41 | 10.2 |
|  | District town | 107 | 26.6 |
|  | Dhaka | 36 | 9.0 |
|  | Divisional district town | 73 | 18.2 |
| HSC- Higher Secondary School Certificate |  |  |  |

### Knowledge and its factors

Participants’ Overall correct answer rate was 89.6% [Mean Score (MS) ± Standard Deviation (SD) = 10.75±1.20 out of 12] while the correct answer rate was 65% to 100%, indicating participants had appreciable knowledge on covid-19. 84.6% of participants had more accurate knowledge while only 15.4% had less adequate knowledge. The distribution of responses for each question of the knowledge scale was presented in (Table 02). Mann-Whitney and Kruskal-Wallis tests showed that knowledge scores do not vary with no socio-demographic variables included in our study, indicating all the groups of a variable had quite similar knowledge on covid-19 disease (Table 04). Multiple logistic regression analysis identified attitude and fear as significant factors that can predict if an individual would have more accurate knowledge on covid-19 rather than having less accurate knowledge (Table 05).

**Table 02.** Participants’ knowledge, attitude, and practice towards covid-19 (N = 402) (Distribution of Responses)

| Knowledge Questions (Correct Answer) | Right<br>n<br>(%) | Wrong<br>n<br>(%) | Not Sure<br>n<br>(%) |
| --- | --- | --- | --- |
| Covid-19 disease is deadly but preventable as well as curable. (Right) | 381<br>(94.8) | 8<br>(2.0) | 13<br>(3.2) |
| Fever, fatigue, dry cough, and breathing difficulty are the main clinical symptoms of covid-19. (Right) | 376<br>(93.5) | 12<br>(3.0) | 14<br>(3.5) |
| Elderly people having chronic illnesses and other complications are more likely to be seriously affected. (Right) | 390<br>(97.1) | 2<br>(0.5) | 10<br>(2.5) |
| This disease can be prevented with antibiotics and other medications. (Wrong) | 95<br>(23.6) | 198<br>(48.3) | 109<br>(27.1) |
| A person with covid-19 having no symptoms (fever) cannot spread the virus to others. (Wrong) | 36<br>(9.0) | 326<br>(81.1) | 40<br>(10.0) |
| Coronavirus can spread via respiratory droplets (from coughing, sneezing) of infected people. (Right) | 389<br>(96.8) | 9<br>(2.2) | 4<br>(1.0) |
| Taking measures is not necessary for children and young adults to prevent infections by the coronavirus. (Wrong) | 7<br>(1.7) | 379<br>(94.3) | 16<br>(4.0) |
| Isolation and treatment of people with covid-19 are effective ways to reduce the spread of the virus. (Right) | 373<br>(92.8) | 14<br>(3.5) | 15<br>(3.7) |
| People who have contact with someone infected with the COVID-19 virus or came from an infected area/country should be immediately isolated for 14 days. (Right) | 393<br>(97.8) | 5<br>(1.2) | 4<br>(1.0) |
| Covering the mouth and nose with a bent elbow or tissue paper or handkerchief when coughing or sneezing can reduce the spread of this virus. (Right) | 377<br>(93.8) | 16<br>(4.0) | 9<br>(2.2) |
| This disease could be asymptomatic. (Right) | 344<br>(85.6) | 26<br>(6.5) | 32<br>(8.0) |
| Individuals should avoid going to crowded places such as markets and public transportations to prevent infection. (Right) | 394<br>(98.0) | 6<br>(1.5) | 2<br>(0.5) |

**Table 02 (Continued)**
| Attitude Questions | Yes<br>n<br>(%) | No<br>n<br>(%) | Maybe/Not<br>Possible for<br>work<br>n<br>(%) |
| --- | --- | --- | --- |
| Do you like to stay at home for a certain period (2 weeks) to prevent coronavirus spread if the government will order so? ** | 345<br>(85.8) | 25<br>(6.2) | 32<br>(8.0) |
| Do you think that social distancing (e.g. stay 1-2 m apart, avoid crowds, etc.) can prevent the spread of this virus? | 338<br>(84.1) | 9<br>(2.2) | 51<br>(13.7) |
| Do you agree that we should cancel business/ recreational trips at this time? | 266<br>(66.2) | 80<br>(19.9) | 56<br>(13.9) |
| Do you believe that working from home can help to control covid-19? | 298<br>(74.1) | 32<br>(8.0) | 73<br>(17.9) |
| Do you agree that government should have taken preventive measures when new variants were first reported in Bangladesh? | 383<br>(95.3) | 3<br>(0.7) | 16<br>(4.0) |
| Do you think health education can play an important role in covid-19 prevention? | 376<br>(93.5) | 7<br>(1.7) | 19<br>(4.7) |
| Do you agree that COVID-19 will finally be successfully controlled? | 176<br>(43.8) | 26<br>(6.5) | 200<br>(49.8) |
| Do you have confidence that Bangladesh can win the battle against the COVID-19 disease? | 139<br>(34.6) | 51<br>(12.7) | 212<br>(52.7) |

**Table 02 (Continued)**
| Practice Questions | Yes<br>n<br>(%) | No<br>n<br>(%) | Sometimes<br>n<br>(%) |
| --- | --- | --- | --- |
| Do you wash hands with water and soap frequently? | 251<br>(62.4) | 32<br>(8.0) | 119<br>(29.6) |
| Do you always use a mask? | 322<br>(80.1) | 20<br>(5.0) | 60<br>(14.9) |
| Do you maintain the rules of using a mask? | 320<br>(79.6) | 23<br>(5.7) | 59<br>(14.7) |
| Do you maintain social distance (or home quarantine)? | 241<br>(60.0) | 69<br>(17.2) | 92<br>(22.9) |
| Do you use tissues or hanker chips during coughing/sneezing? | 240<br>(59.7) | 92<br>(22.9) | 70<br>(17.4) |
| Do you eat healthy food or maintain a healthy lifestyle focusing on outbreaks? | 269<br>(66.9) | 42<br>(10.4) | 91<br>(22.6) |
| Do you avoid public transports (like buses, trains, planes, etc.)? | 183<br>(45.5) | 127<br>(31.6) | 92<br>(22.9) |
| Do you avoid handshaking, hugging when you meet with your friends? | 238<br>(59.2) | 84<br>(20.9) | 80<br>(19.9) |
| ** This question had “Not possible for work” instead of “Maybe” in its three options. n- Number of respondents, %- Percentage of respondents, N- Total number of respondents |  |  |  |

**Table 03.** Fear related to covid-19 among respondents (N= 402) (Distribution of responses).

| Fear Questions | Strongly Disagree<br>n<br>(%) | Disagree<br>n<br>(%) | Neutral<br>n<br>(%) | Agree<br>n<br>(%) | Strongly Agree<br>n<br>(%) |
| --- | --- | --- | --- | --- | --- |
| I am most afraid of coronavirus-19. | 48<br>(11.9) | 106<br>(26.4) | 127<br>(31.6) | 92<br>(22.9) | 29<br>(7.2) |
| It makes me uncomfortable to think about coronavirus-19. | 38<br>(9.5) | 126<br>(31.3) | 97<br>(24.1) | 114<br>(28.4) | 27<br>(6.7) |
| My hands become clammy when I think about coronavirus-19. | 127<br>(31.6) | 228<br>(56.7) | 29<br>(7.2) | 15<br>(3.7) | 3<br>(0.7) |
| I am afraid of losing my life because of coronavirus-19. | 53<br>(13.2) | 143<br>(35.6) | 102<br>(25.4) | 84<br>(20.9) | 20<br>(5.0) |
| When watching news and stories about coronavirus-19 on social media, I become nervous or anxious. | 50<br>(12.4) | 126<br>(31.3) | 106<br>(26.4) | 88<br>(21.9) | 32<br>(8.0) |
| I cannot sleep because I'm worried about getting coronavirus-19. | 140<br>(34.8) | 216<br>(53.7) | 32<br>(8.0) | 13<br>(3.2) | 1<br>(0.2) |
| My heart races or palpitates when I think about getting coronavirus-19. | 135<br>(33.6) | 190<br>(47.3) | 51<br>(12.7) | 24<br>(6.0) | 2<br>(0.05) |

n- Number of respondents, %- Percentage of respondents, N- Total number of respondents

**Table 04.**
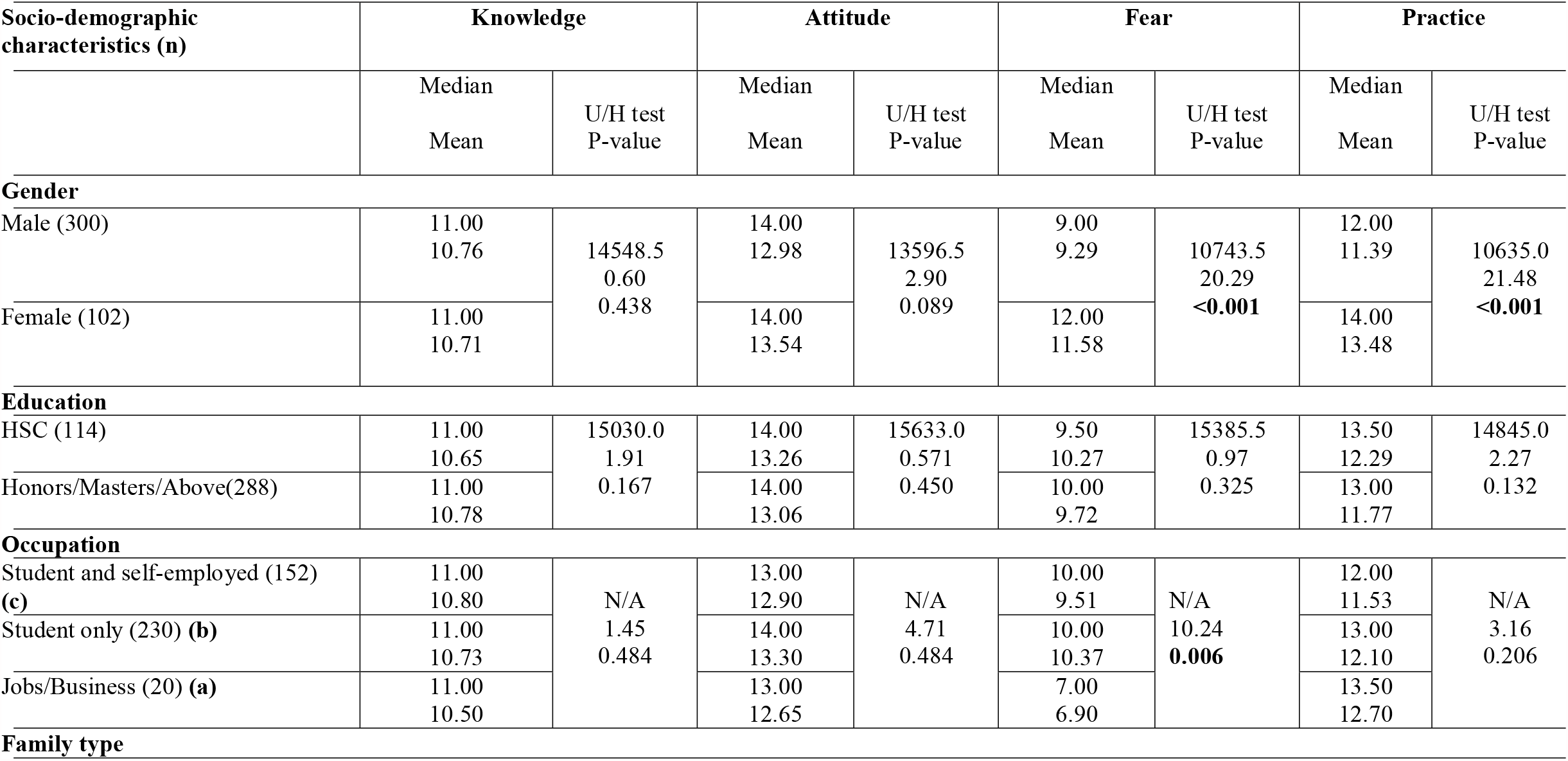
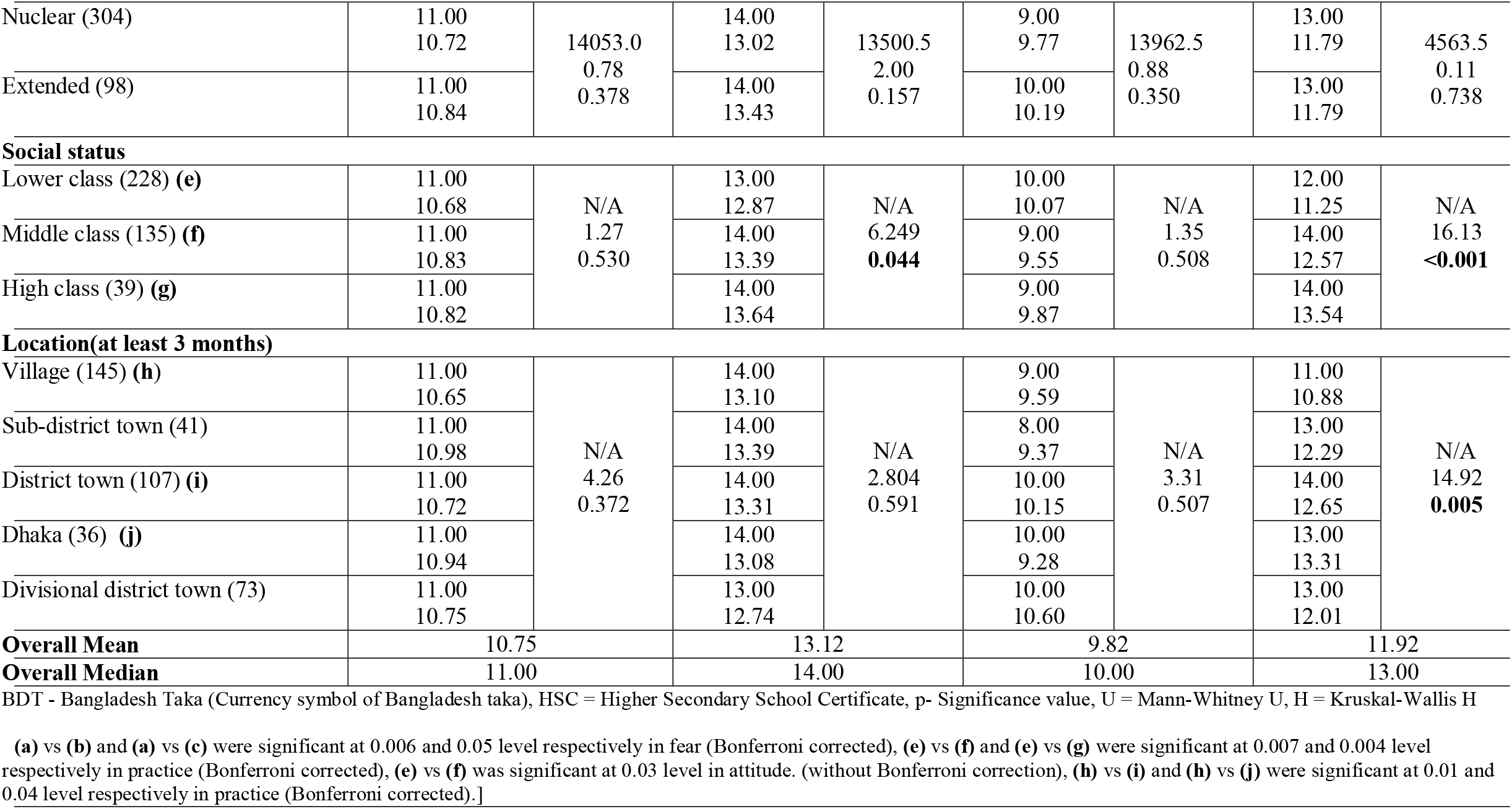
Association of socio-demographic characteristics with knowledge, attitude, fear, and Practice score.

**Table 05.**
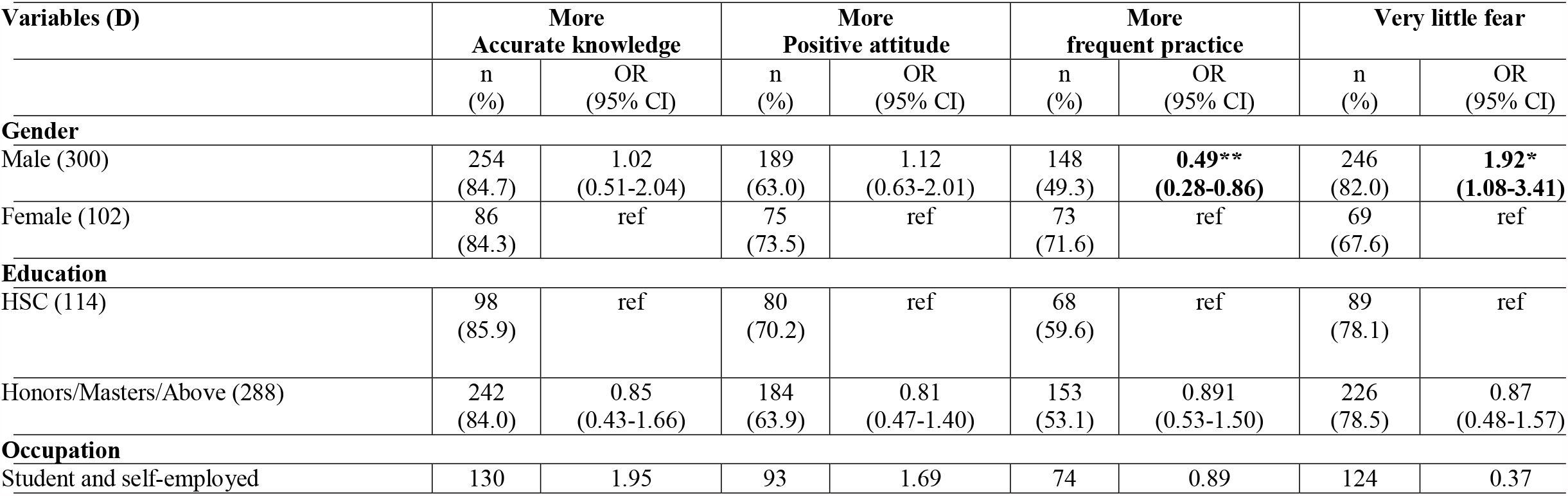
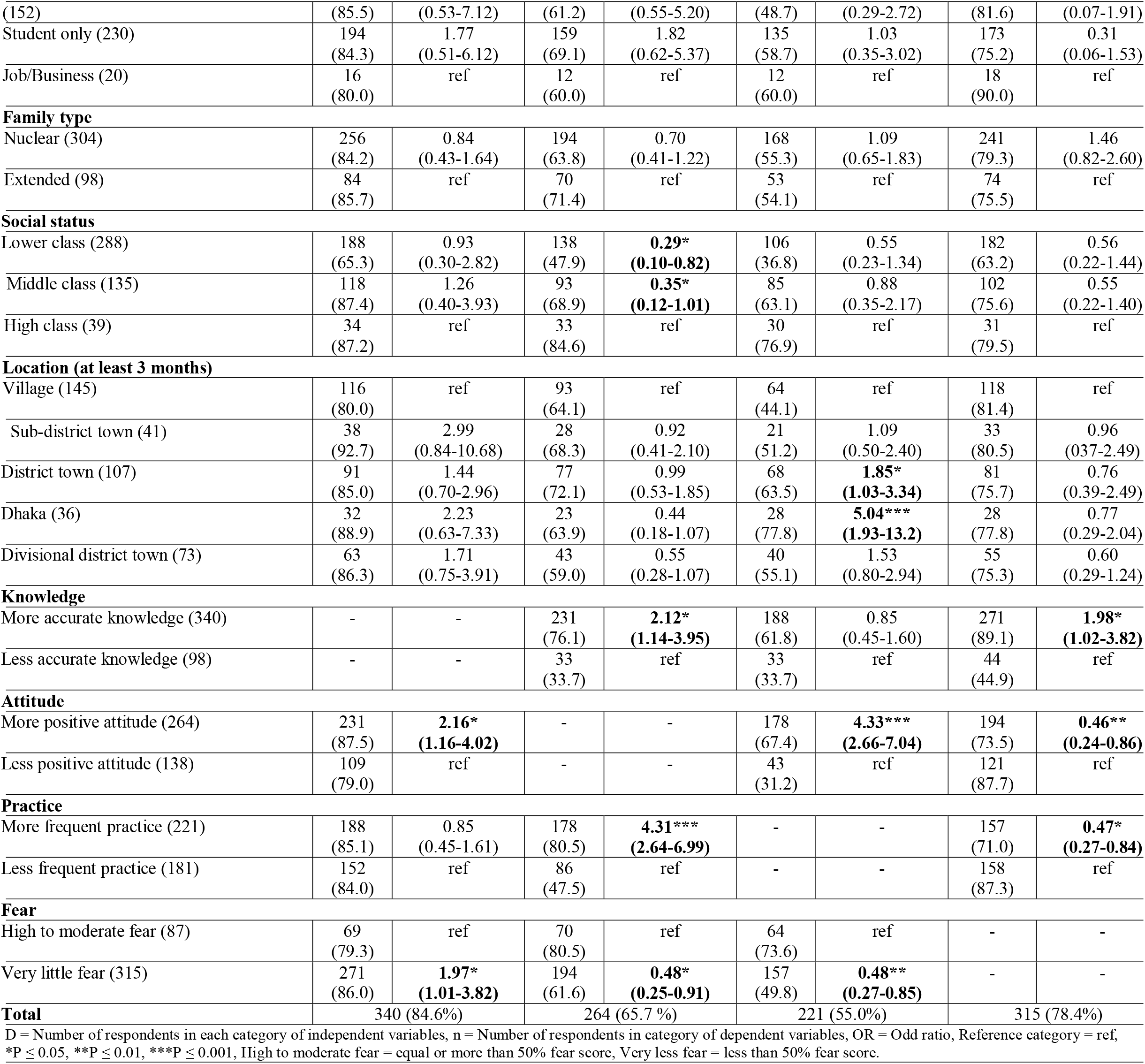
Results of multiple logistic regression on factors associated with knowledge, attitude, practice, and fear. (Adjusted model)

### Attitude and its factors

On average, participants had an 82% score on the attitude scale (MS ± SD = 13.12 ± 2.21 out of 16). Approximately 66% of participants had a more positive attitude. Only 34.6% of participants had confidence that Bangladesh can win the battle against covid-19 and only 43.8% agreed that covid-19 will finally be controlled. The distribution of responses for each question of the attitude scale was presented in (Table 02). Mann-Whitney and Kruskal-Wallis tests showed attitude scores vary with only social status (P< 0.05). It was found that participants who came from a lower-class family had significantly less attitude than those who came from a middle-class family (P < 0.05) while other combinations were statistically insignificant (Lower class vs High class; Middle class vs High class) (Table 04). Multiple logistic regression analysis identified social status, knowledge, practice, fear, as significant factors that can predict if an individual would have a more positive attitude rather than a less positive attitude. It was found that participants who came of a lower class family were 71% less likely to have more positive attitude than those who came of a high-class family (OR = 0.29, 95% CI = 0.10-0.82, P < 0.05) and participants came of a lower class family were 65% less likely to have more positive attitude than those came of a high-class family (OR = 0.35, 95% CI = 0.12-1.01, P < 0.05) (Table 05).

### Practice and its factors

On average, participants had a 74.5% score on the practice scale (MS ± SD = 11.92 ± 3.77 out of 16). 55% of participants had more frequent practice. The distribution of responses for each question of the practice scale was presented in (Table 02). Mann-Whitney and Kruskal-Wallis tests uncovered that Practice scores vary with gender (P < 0.001), social status (P < 0.001), and location (P < 0.01). It was found that females had significantly higher practice score than males (P < 0.001); Participants came of both higher class family (P < 0.01) and middle-class family (P < 0.01) had significantly higher practice score than those came of a lower class family; Participants living in Dhaka (P < 0.05) and in any district town (P = 0.01) for at least three months had significantly higher score on practice scale than those living in a village for the same duration of time (Table 04). Multiple logistic regression analysis identified gender, location, attitude, fear, as significant factors that can predict if an individual would have more frequent practice rather than less frequent practice. It was found that males were less likely to have more frequent practice than females by 51% (OR = 0.49, 95% CI = 0.28-0.86, P ≤ 0.01). Participants living in any district town were 85% more likely to have more frequent practice than those living in a village (OR = 1.85, 95% CI = 1.03-3.34, P ≤ 0.05) while Participants living in Dhaka were 5.04 times more likely to have more frequent practice than those living in a village (OR = 5.04, 95% CI = 1.93-13.2, P ≤ 0.001) (Table 05).

### Fear and its factors

On average, participants had a 35.5% fear score (MS ± SD = 9.87 ± 4.78 out of 28). 78.4% (315) participants had less fear, 19.7% (79) had moderate fear, and only 2.0% (8) participants had high fear. And 78.4% (315) participants had very less fear (less than 50% fear score) while only 21.6% (87) participants had high to moderate fear (equal or more than 50% fear score). The distribution of responses of all fear-related questions was presented in (Table 03). Mann-Whitney and Kruskal-Wallis tests unfolded that fear score among participants varies with gender and occupation. It was reported that females had a significantly high fear score than males (P < 0.001); Participants doing job or business had significantly low fear than both those who are student only (P = 0.05) and those who are students as well as self-employed (P < 0.01) (Table 04). Multiple logistic regression analysis detected gender, knowledge, attitude, practice, as significant factors that can predict whether an individual would have very little fear rather than high to moderate fear. It was found that male participants were 92% more likely to have very less fear than female participants (OR = 1.92, 95% CI = 1.08-3.41, P < 0.05) (Table 05).

### Relationship among knowledge, attitude, practice, and fear

Spearman rank-order correlation tests explored that knowledge score is significantly correlated with attitude score (P < 0.01) although it was insignificantly correlated with other scores (practice and fear score). However, attitude score was correlated with both practice score (P < 0.001) and fear score (P < 0.001). Along with attitude score, practice score and fear score was correlated with each other (P < 0.01) (Table 06). Multiple logistic regression analysis uncovered that participants who had more accurate knowledge were 2.12 times more likely to have a more positive attitude (OR = 2.12, 95% CI = 1.14-3.95, P < 0.05) and 1.98 times more likely to have very little fear (OR = 1.98, 95% CI = 1.02-3.82, P < 0.05) than those who had less accurate knowledge; Participants who had a more positive attitude were 4.33 times more likely to have more frequent practice than those who had less positive attitude (OR = 4.33, 95% CI = 2.66-7.04, P < 0.001). Moreover, participants who had very less fear were 52% less likely to have more positive attitude (OR = 0.48, 95% CI = 0.25-0.91, P < 0.05) at the same time 52% less likely to have more frequent practice (OR = 0.48, 95% CI = 0.27-0.85, P < 0.05) than those who had high to moderate fear (Table 05).

**Table 06.** Results of spearman’s rank-order correlation among knowledge, attitude, practice, and fear score.

|  |  |  |  |  |
| --- | --- | --- | --- | --- |
| **Table 06** |  |  |  |  |
|  | **1** | **2** | **3** | **4** |
| 1. Knowledge | 1 |  |  |  |
| 2. Attitude | 0.139<sup>a</sup><br>0.005<sup>b</sup> | 1 |  |  |
| 3. Practice | 0.046<sup>a</sup><br>0.357<sup>b</sup> | 0.358<sup>a</sup><br>< 0.001<sup>b</sup> | 1 |  |
| 4. Fear | -0.049<sup>a</sup><br>0.329<sup>b</sup> | 0.243<sup>a</sup><br>< 0.001<sup>b</sup> | 0.138<sup>a</sup><br>0.006<sup>b</sup> | 1 |

a = Correlation coefficient, b = Significance level (Two tailed)

### Possible reasons of an individual behind failing to maintain all safety rules stipulated for covid-19 prevention

When asked to observe participants’ thoughts as to whether they are maintaining safety rules for covid-19, 25.6% (103) participants assured that they are maintaining safety rule completely while 74.4% (299) reported “No” and mentioned their possible reasons why they were detached from maintaining safety rules properly. Among those who mentioned “No”, 57.5% participants mentioned they cannot maintain a social distance because they need to go out for many reasons and need to use public transports where social distance maintaining is not possible while 44.8% participants mentioned they need to go market for daily groceries or many things where they cannot maintain social distance. 29.1% of participants think of themselves as social butterflies and meet with many friends while they do not maintain social distance. 22.4% of participants reported that they can’t wash their hands regularly while 33.1% mentioned that very few times a day, they wash their hands as recommended by WHO. The further analysis explored that among participants who mentioned they wash their hands regularly, 23.3% do not wash their hands maintaining rules recommended by WHO while 76.7% wash their hands regularly as recommended by WHO and among those who mentioned they do not wash their hands regularly, 32.8% maintain rules recommended by WHO when they wash their hands. However, 37.5% of participants don’t use hand sanitizers when staying outside the home and 23.4% of participants have bad habits of touching eyes, nose, mouth, frequently. 21.4% of participants reported that they have not seen anyone from their relatives or relatives of relatives or neighbors got infected by a coronavirus and that is why they do not give importance to maintaining safety rules of covid-19. (Figure 02) depicted all the other reasons graphically with the frequency of responses for each reason and (Table 07) tabulated all the supportive information.

**Table 07.** Possible reasons why people cannot maintain safety rules of covid-19 completely.

| **Table 07** |
| --- |
| *Possible reasons why people cannot maintain safety rules of covid-19 completely.* |
| ▪ Beliefs that I will not be infected by the coronavirus. (C01) |
| ▪ Things related to corona seem to me as media-created rumors. (C02) |
| ▪ No one having been identified as corona virus-positive yet from my relatives or relatives of relatives or neighbors makes maintaining safety rules less important to me. (C03) |
| ▪ I am less panicked about coronavirus. (C04) |
| ▪ I have no clear conception about safety rules for covid-19. (C05) |
| ▪ It is not always possible for me to wash hands coming from outsides. (C06) |
| ▪ Very few times in a day, hands are washed up to the elbow for (20-30) min as recommended by WHO. (C07) |
| ▪ I do not use hand sanitizer when staying outside. (C08) |
| ▪ Trying but I can't give up the bad habit of touching eyes, face, nose frequently. (C09) |
| ▪ I am to go to the market often for daily groceries or other things where maintaining social distance is not possible. (C10) |
| ▪ I am to go out for many reasons and use public transports where maintaining social distance is not possible. (C11) |
| ▪ I am a social butterfly and hang out with friends very often while social distance is not maintained. (C12) |
| ▪ I don't maintain social distance thinking about the benefit of maintaining social distance when people around me don't maintain social distance. (C13) |
| ▪ I don't use medical masks because of having any confidence in a normal medical mask (5 BDT) as well as no |
| money for buying expensive masks. (C14) |
| ▪ When wearing a mask, I feel very uneasy and it's very hard to stay with. (C15) |
| ▪ I don't use a mask because it looks ugly. (C16) |
| ▪ I don't know any reason. (C17) |
| BDT-Bangladesh Taka (Currency symbol of Bangladeshi taka) |

## Discussion

To fulfill the goals of this present study, several statistical analyses had been conducted and their outcomes had been presented sequentially in the result section. In this section, these outcomes have been discussed collaboratively with other previously published studies.

### Knowledge level of Participants

The findings showed that participants’ overall correct answer rate was 89.6%, indicating participants had appreciable Knowledge on covid-19. This outcome was consistent with some previous studies conducted last year (2020) in Bangladesh (80.8%) (17), China (90%) (18), in Malaysia (80.5%) (19), in Saudi Arabia (81.6%) (20), in Iran (90% & 85%) (21), in Nepal (60% to 98.7%) (22). Moreover, 84.6% of participants had more accurate knowledge which was larger compared to that reported by some previous Bangladeshi investigations (6–10), indicating knowledge level has elevated among Bangladeshis.

Our study reported that knowledge scores do not significantly vary with gender, education, occupation, location, social status, family type, which was inconsistent with some studies (6–10,18,23).

### Attitude level of Participants

Approximately 66% of participants had a more positive attitude towards covid-19, which was commensurate with (7). But when compared to the other two Bangladeshi articles, it seems attitude level among people has alleviated to some extent (9,10). Compared to overseas investigations, our finding was consistent with a study from Pakistan (65.4%) (11), while inconsistent with studies from Saudi Arabia (95%) (24), China (73.8%) (13). Our findings suggested that Bangladeshi adults might have lost their confidence towards controlling the rapid escalation of coronavirus as only 34.6% of participants took it confidently that Bangladesh can win the battle against covid-19 and concomitantly, only 43.8% of participants were confident that covid-19 will finally be controlled. Last year, Rabbani et al., (2020) reported that 55.3% of their participants were confident as to Bangladesh would win the battle against covid-19 which was also consistent with Kundu et al., n.d. (2020), and 68.5% were confident that covid-19 would be finally controlled which was somewhat lower compared to Banik et al., (2020) (8,9,25). On the other hand, a Malaysian, Chinese, Saudi Arabian, Indonesian, Nepali article published last year (2020) reported that they respectively obtained 83.1%, 90.8%, 94%, 94.3%, and 71.5% positive responses regarding whether covid-19 would successfully be controlled, and 95.9%, 97.1%, 97%, 95.5%, and 80% positive response regarding whether their country would be able to win the battle against covid-19 (18– 20,23,26).On the whole, it is obtrusive that attitude level among the populace has noticeably subsided. The reason behind obtaining such ominous outcomes might be the long duration of uncertainties which has been appeared due to emerging of new variants of SARS-COV-2 with the function of time.

Moreover, our study explored that attitude levels significantly vary with only the social status of the participants, not with other socio-demographic variables included in our scrutiny, which was somewhat inconsistent with several studies. (6,7,9).

### Fear level of Participants

On average, participants had a 35.5% fear score which was lower compared to M. A. Hossain et al., (2020) (27). Besides, only 2% of participants had very high fear while 78.4% had very less fear. These outcomes indicate that Bangladeshi people are no longer panicked. Furthermore, the fear score was significantly associated with the gender and occupation of participants. Females had significantly high fear than males and participants doing a job or business had significantly low fear than both those who are student only and those who are student as well as self-employed. M. A. Hossain et al., (2020) reported that the fear score significantly differs with gender, education, and geography, when female, urban-dwelling, and higher educated participants had high fear, which was partially similar to our findings (27).

### Practice level of Participants

On average, participants had a 74.5% practice score and 55% participants had more frequent practice, which was consistent with some studies from Bangladesh (6,7,9,25). So, our study indicates that the preventive practice level remains similar to what was in last year. However, compared to a study from Bangladesh (24%) (8) and from Pakistan (36.5%) (11), our participants had more frequent practice but seems to have much lower frequent practice when compared to studies from Saudi Arab (81% & 81.9%) (12,24) and South Korea (87.9%) (13). Indeed, our study found that more frequent practice is greatly influenced by gender and location. However, the more frequent practice could also be influenced by other socio-demographic factors such as education, occupation, age, family income, etc. (7– 10).

### Relationship among knowledge, attitude, practice, and fear

The finding showed knowledge has an indirect effect on preventive practice mediated by attitude as knowledge has a positive impact on attitude and concomitantly, an attitude has a positive impact on preventive practice, indicating the more knowledge on covid-19, the more attitude towards covid-19, and the more frequent practice towards covid-19 prevention. Similar outcomes were reported in some other studies (8,10,14,24,28,29). Moreover, our study uncovered that knowledge has a significant inverse effect on fear, indicating the more accurate knowledge, the less fear. This outcome was parallel with some other studies (17,24,27,30). Our study also revealed that like attitude, fear has also a positive impact on preventive practice, indicating the more fear, the more preventive practice. A similar outcome was reported by (27,31). Ostensibly, this outcome could seem controversial but it represents the significant influence of fear on changing someone’s behavior or attitude towards preventive practice. Our study also substantiates the immediate statement reporting that fear has a positive and mysteriously same impact on attitude as was on preventive practice. However, we cannot expect high fear among the populace to have more practice because fear has some shortcomings too. For instance, Covid-19 fear is significantly associated with anxiety and psychological distress could also trigger stress and mental illness (30). Besides, there is an instance of suicidal death due to the fear of covid-19 in Bangladesh (27,32). (Figure 03) depicted all the factors and their impact on preventive practice.

**Fig 3.**
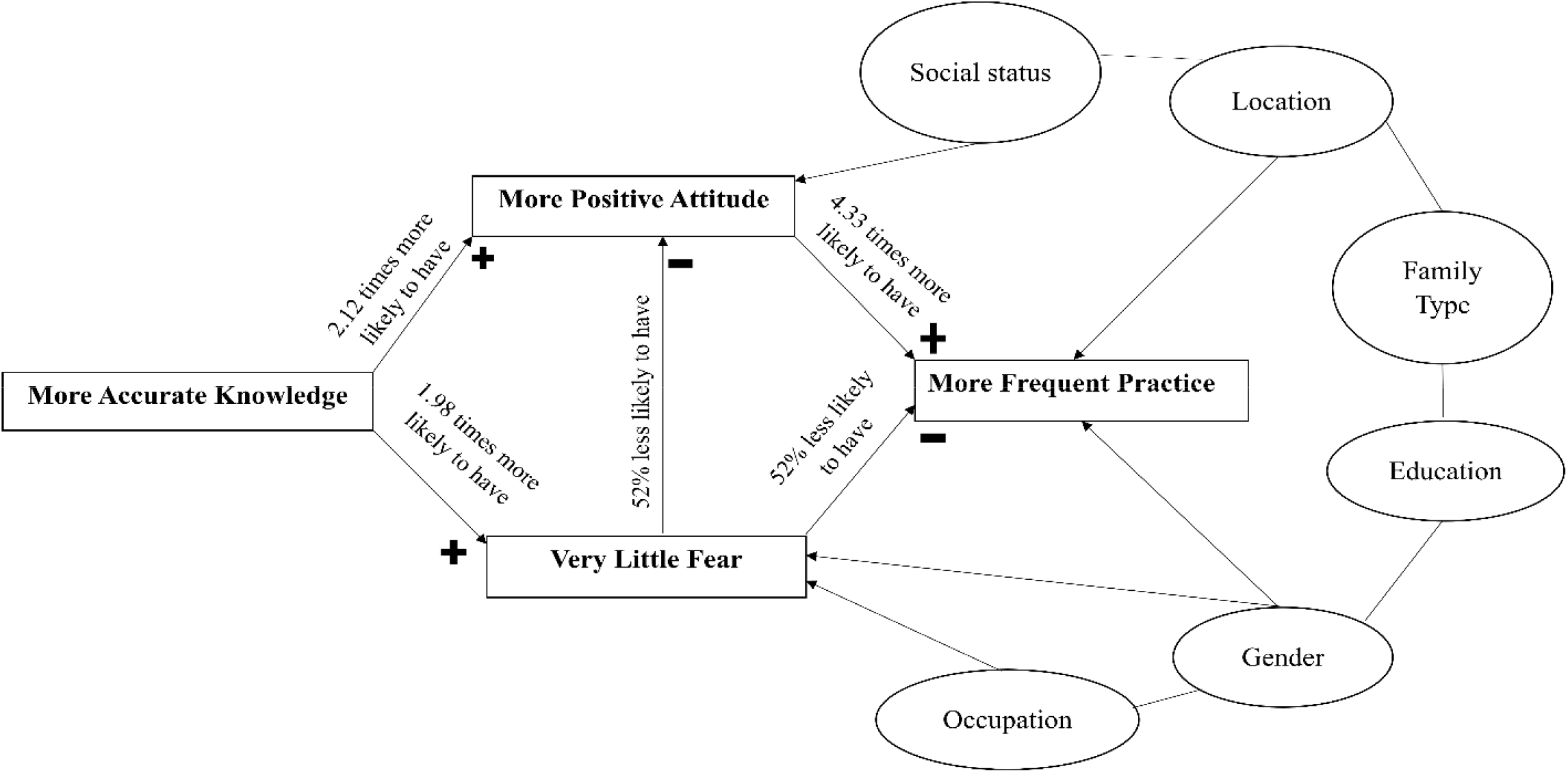
Diagram showing the direct and indirect impact of knowledge, attitude, fear, and socio-demographic factors (Ellipse) on preventive practice.

Moreover, Our study reported that the majority of the participants thought they cannot adhere to preventive measures because of not maintaining social distance properly, followed by reluctance in hand washing as recommended by WHO. We had no sufficient data to support these reasons.

### Limitations, Strength, and Recommendation

Our study had several limitations. First, our study sample could not reflect all populations throughout the country. Second, our study could not include participants of all ages. Third, the study sample size was comparatively small. Fourth, our study followed a web-based cross-sectional study that excluded underprivileged people especially those who were not accessible to internet-based facilities. Five, we had to conduct comparatively less sensitive statistical analysis as our data were not normally distributed. As to the strength of our study, to the best of our knowledge, this is the first study that attempted to find out possible reasons why people are deflected from maintaining preventive measures. Future investigations should follow both community-based and web-based cross-sectional studies, collecting data from all sectors, all divisions, attempting to find out the most probable reasons behind failure to maintain all preventive measures for covid-19 prevention in a more structured way.

## Conclusion

Our study attempted to bring up the present knowledge, attitude, practice, and fear level of Bangladeshi people. According to our study, participants had appreciable knowledge and very little fear but disappointedly had average attitude and practice. Besides, Participants had a lack of confidence on Bangladesh would win the battle against covid-19. Therefore, it is time for public health policymakers to be more focused to scale up people’s confidence and attitude towards covid-19. To do so, they must be more dogmatic to develop rules and regulations and more austere in implementing and maintaining those to combat any new onslaught of the deadly coronavirus. Besides, providing public health educations arranging viable public health campaigns, promoting accurate information regarding covid-19, counseling general people with regards to preventive practices, must be continued till the end of this unprecedented condition.

## Data Availability

For data and material requests, please email the corresponding author.

## Acknowledgments

The authors acknowledge all the participants who participated in the study and all the volunteers who helped in data collection. The authors are also grateful to Dr. Md. Abdul Rafi for his support in the overall improvement of the manuscript.

## Author contributions

**Conceptualization:** Tahsin Ahmed Rupok, Sunandan Dey

**Data curation:** Tahsin Ahmed Rupok, Rashni Agarwala, Md. Nurnobi Islam, Sunandan Dey, Bayezid Bostami.

**Formal Analysis:** Tahsin Ahmed Rupok, Rashni Agarwala.

**Investigation:** Tahsin Ahmed Rupok, Rashni Agarwala, Md. Nurnobi Islam, Sunandan Dey.

**Methodology:** Tahsin Ahmed Rupok, Bayezid Bostami.

**Project Administration:** Tahsin Ahmed Rupok, Sunandan Dey.

**Supervision:** Tahsin Ahmed Rupok, Sunandan Dey.

**Writing – original draft:** Tahsin Ahmed Rupok.

**Writing – review & editing:** Tahsin Ahmed Rupok, Md. Nurnobi Islam, Sunandan Dey, Rashni Agarwala, Bayezid Bostami.

